# Evaluation of Type 2 Diabetes Like Subtypes in Gestational Diabetes

**DOI:** 10.64898/2026.07.29.26359198

**Authors:** Luma Srour, Nayra M. Al-Thani, Eleni Fthenou, Omar M. E. Albagha, Nady El Hajj

## Abstract

**Introduction:** Gestational diabetes mellitus (GDM) is a condition characterized by glucose intolerance that is first identified during pregnancy and typically resolves after childbirth. This condition can lead to various complications, both prenatal and postnatal, including type 2 diabetes (T2D). However, not all women with GDM progress to T2D, and the molecular mechanisms underlying this heterogeneity remain poorly understood. Here, we hypothesized that GDM participants could be clustered into T2D-like subtypes.

**Methods:** To derive T2D-like subtypes, we applied K-means clustering to GDM participants using the predefined T2D subtype cluster centers established in the QPHI cohort. Differential methylation analysis was performed, and the top-ranked sites were used for further analysis. Additionally, we estimated system-specific age acceleration and its association with subtype-specific complications.

**Results:** Our study demonstrated the effectiveness of the novel clustering approach, originally developed for T2D, in GDM. Additionally, the exploratory analysis of the top-ranked CpG sites suggested potential subtype-related methylation patterns. The identified pathways also suggested overlapping molecular mechanisms underlying both GDM and T2D.

**Conclusion:** These findings support the feasibility of classifying GDM into T2D-like clinical subtypes and suggest potential epigenetic differences that warrant validation in larger longitudinal cohorts.

## Introduction

Gestational diabetes mellitus (GDM) is one of the most common metabolic complications during pregnancy. It is characterized by glucose intolerance first recognized during pregnancy that usually resolves after delivery [1]. Women with GDM are generally advised to undergo regular postpartum screening for type 2 diabetes (T2D), as they have an approximately 8.3-fold higher risk compared with women without GDM [2, 3]. This risk becomes even higher with age, BMI, and among Asian women [2]. Despite effective disease management through lifestyle changes or medication, physiological dysregulation associated with gestational hyperglycemia contributes to an increased risk of future metabolic diseases, including T2D [4].

Previous studies have indicated epidemiological, physiological, and partial genetic similarity between T2D and GDM [1, 5]. Similar features have been identified in both diseases, including lipid deposition in the muscle and liver, increased adipose tissue macrophages, and the secretion of pro-inflammatory cytokines, which play a major role in insulin impairment and inhibition of insulin release from β-cells [6]. Notably, T2D is a highly heterogeneous disease; thus, considerable variation exists among participants in their clinical characteristics, disease progression, complication development, and treatment response [7]. Due to this variation, a novel model was proposed to categorize T2D into four subtypes: severe insulin-deficient diabetes (SIDD), severe insulin-resistant diabetes (SIRD), mild obesity-related diabetes (MOD), and mild age-related diabetes (MARD). This model utilized several measures, including age, BMI, hemoglobin A1c (HbA1C), homeostatic model assessment of β-cell function (HOMA2-B), and insulin resistance (HOMA2-IR) [7]. Clustering of T2D participants using those clinical variables enables personalized treatment strategies [8]. Similarly, GDM is considered a highly heterogeneous disease [9, 10]. Despite the considerable variability observed among women with GDM, all are treated similarly. The initial stage of treatment focuses on lifestyle changes, while the second stage involves medical intervention if the glycaemic goals are not achieved [9, 11]. Given the similarities between GDM and T2D, it is essential to stratify GDM participants by diabetes-related parameters. Clustering GDM into subgroups may help assess disease severity and refine treatment selection [9]. However, it remains unclear whether the novel T2D clustering model will be effective for GDM.

GDM is considered a complex condition influenced by genetic, epigenetic, and environmental factors [12]. Epigenetic mechanisms, including DNA methylation, have been implicated in the pathogenesis of GDM and are thought to influence maternal metabolic adaptations during pregnancy [13, 14]. Growing evidence suggests that gestational hyperglycaemia is associated with alterations in DNA methylation, some of which may persist beyond pregnancy [4]. These epigenetic changes may reflect differences in the underlying biological mechanisms contributing to GDM [15]. However, whether DNA methylation changes during pregnancy capture the molecular heterogeneity underlying distinct GDM subtypes remains unexplored.

In this study, we used the Ahlqvist *et al.* [7] clustering method to assess its applicability for identifying GDM subtypes and for characterizing GDM clinical heterogeneity. We hypothesized that GDM participants could be clustered into T2D-like subtypes, thereby facilitating the evaluation of GDM heterogeneity. We examined longitudinal changes in GDM subtypes between the second (T2) and third (T3) trimesters. Using longitudinal data from T2 and T3, we additionally aimed to identify epigenetic changes that persisted across both trimesters and were associated with each subtype.

## Materials and Methods

### Study Cohort (Qatar Birth Cohort)

A total of 50 individuals with GDM were selected from our previous study on gestational diabetes [16]. These represent a subset of the Qatar Birth Cohort (QBiC) study, recruited at Qatar Precision Health Institute (QPHI) [17]. As described previously, these participants were self-reported pregnant women with GDM who were ≥ 18 years old. Participants with longitudinal data available at both T2 (14–28 weeks) and T3 (29–40 weeks) were selected from the original cohort. The data included clinical parameters, biochemical measurements, and electronic questionnaires collected at each time point at Hamad Medical Corporation (HMC). Informed consent was obtained from all pregnant women who participated in this study in accordance with the Declaration of Helsinki. This study was approved by the institutional review boards of QPHI (Ex-2022-QF-QBB-RES-ACC-00100-0203) and Hamad Bin Khalifa University (QBRI-IRB 2021-09-107).

### Clustering Analysis

We used the established, predefined T2D subtype cluster centers for the QPHI cohort [18] to derive T2D-like subtypes among GDM participants in the QBiC cohort. We conducted K-means clustering using the factoextra library in R (v 4.4.2). The clustering was performed using the clinical variables HbA1C, BMI, age, HOMA2-%B, and HOMA2-IR, with standardized values (mean of 0 and standard deviation of 1) for all GDM participants, separately for each trimester (T2 and T3). Prior to the analysis, HOMA2-%B and HOMA2-IR were calculated using the HOMA2 calculator [19] based on the C-peptide levels. Following the subclassification of GDM participants, a Sankey diagram was constructed to illustrate the transition in GDM cluster assignment for individuals from T2 to T3.

### Differential Methylation and Downstream Analysis

DNA methylation profiling for the study cohort was conducted according to the manufacturer’s instructions using the Infinium MethylationEPIC v2.0 BeadChip. Raw intensity data files (IDATs) were chosen for participants with longitudinal data at both time points, T2 and T3. Using the Rnbeads package in R, preprocessing, normalization via the beta-mixture quantile normalization method (BMIQ), and blood cell estimation using the FlowSorted.Blood.EPIC package was performed as previously described [16]. Rnbeads removed 9981 probes that overlapped with known SNPs, in addition to 20744 probes that were filtered out via the GreedyCut algorithm. Differential methylation analysis was also conducted at both the site and region levels (including promoters, CpG islands, and tiling in 5-kb windows) for each subtype compared to all others. Due to a lack of methylation data for the T2 samples clustered into the SIRD subtype, we excluded the SIRD subtype from the differential methylation analysis and subsequent downstream analyses. Thus, the following comparisons were performed: MARD vs. All, MOD vs. All, and SIDD vs. All. Differential methylation analysis was performed using M-values (logit-transformed beta values) and linear modeling implemented in the R package limma. We adjusted for several covariates in the linear modeling, including age, BMI, blood cell proportions, and batch effect. The significance of the result was defined by an FDR-adjusted *p-value < 0.05*, while biological significance was also assessed by a Δβ > 2% methylation difference between the subgroup and all other subgroups. When no sites or regions reached significance after FDR correction, they were prioritized using a combined rank based on the mean methylation-level differences, the mean methylation quotient, and the *p-value*.

After conducting the differential methylation analysis, the top 10,000 CpG sites from each comparison at T2 and T3 were identified, since no CpG sites remained significant after FDR correction (FDR-adjusted *p-value* < 0.05). Subtype-specific CpG sites were identified by overlapping the T2 and T3 sites for each comparison. These sites were then used for downstream analysis. In addition, the top 100 combined rank promoters from each comparison at T2 and T3 were identified. Differentially methylated promoters associated with each subtype were identified by overlapping the T2 and T3 regions for each comparison. Subtype-specific CpG sites were used for downstream analysis. Kyoto Encyclopedia of Genes and Genomes (KEGG) pathway and Gene Ontology (GO) enrichment analyses were conducted using the methylGSA package in R (version 4.4.2).

In this study, system-specific age was estimated using the calcSystemsAge function from the methylCIPHER package. This package incorporates several clocks, including the new systems-based methylation clock. This clock can estimate age in multiple physiological systems, including Heart, Lung, Kidney, Liver, Brain, Immune, Inflammatory, Blood, Musculoskeletal, Hormone, and Metabolic [20]. In addition, we estimated biological age using GrimAge and PhenoAge, based on the DNA methylation age (DNAmAge) calculator from the Steve Horvath lab. Epigenetic age acceleration was also computed as the residual of regressing system-specific age and epigenetic age on chronological age, while adjusting for BMI and batch. Furthermore, we estimated the pace of aging in GDM participants using DunedinPACE. DunedinPACE represents a blood DNA methylation biomarker of the pace of biological aging, with values indicating whether an individual is aging faster or slower than the normative rate of one year of aging per year of calendar time. Due to a lack of methylation data for the SIRD subgroup at T2, this subgroup was excluded from the epigenetic age acceleration analysis at T2.

To assess aging across biological systems, we calculated the mean age acceleration for each diabetes subtype (SIDD, SIRD, MOD, and MARD) at two time points (T2 and T3) in R (v 4.4.2). We then generated spider plots to visualize age acceleration for each system across subtypes and to assess consistency over time.

### Shared DNA Methylation Signatures Between GDM and T2D

To identify CpG sites that could trigger the onset of type 2 diabetes (T2D), we conducted differential methylation analysis in a BMI-matched T2D cohort from the same population as our study participants. This cohort represents a subset of the individuals included in the clustering analysis reported by Al-Thani *et al.* in 2025 [18]. This cohort included 48 participants, distributed across T2D subtypes as follows: MARD = 16, MOD = 11, SIDD = 12, and SIRD = 9. The following comparisons were conducted during the methylation analysis in this cohort: MARD vs. All; MOD vs. All; SIDD vs. All; and SIRD vs. All. We identified the top 100 unique CpG sites for each T2D subtype. We compared them with previously identified GDM-associated CpG sites to identify shared methylation markers between GDM and T2D. In addition, we investigated common CpG sites between the discovered GDM-associated sites and those previously identified by Schrader *et al.* as T2D subgroup-unique CpG sites [21].

### Statistical testing

Several clinical characteristics, including age, BMI, HbA1C, HOMA2-%B, and HOMA2-IR, across trimesters and GDM subgroups were compared using GraphPad Prism (version 10.4.0). To assess normality for each variable, the Shapiro-Wilk test was applied, and the test choice was based on the data’s normality. The changes across trimesters were assessed using a paired t-test for BMI and HbA1C. While the changes in HOMA2-%B and HOMA2-IR across trimesters were assessed using the Wilcoxon matched-pairs signed-rank test, depending on the distribution of each variable. A one-way analysis of variance (ANOVA) was performed to evaluate the significance of variables such as Age, BMI, and HOMA2-%B across GDM subgroups at T2, while the Kruskal-Wallis test was used for HOMA2-IR. Similarly, at T3, a one-way ANOVA assessed Age, BMI, and HOMA2-%B, and the Kruskal-Wallis test was applied for HbA1C and HOMA2-IR to determine their significance across GDM subgroups. In addition, we compared system-specific and epigenetic age acceleration (EAA) using GraphPad Prism. A one-way ANOVA was used for most of the EAA, while the Kruskal-Wallis test was used for blood, liver, musculoskeletal, and DunedinPACE at T2. Similarly, the Kruskal-Wallis test was used for heart, hormone, lung, and phenoAge at T3.

We also calculated the changes by subtracting the mean values of each variable at T3 from those at T2 across all transitions, including both stable and switch samples. Stable samples refer to participants who retained the same subtype classification between T2 and T3 (e.g., MARD→MARD). On the other hand, switch samples are participants who changed cluster classification between T2 and T3 (e.g., MARD→SIRD). For this analysis, *p-values* were calculated using paired t-tests for most subtype transitions. However, for the MOD → MARD transition in HbA1c, the MOD → MOD transition in the change of HOMA2-IR, and the MARD → MARD transition in HOMA2-%B, the Wilcoxon signed-rank test was applied due to non-normal distribution of the paired differences. The *p-value* was calculated for transitions with 3 or more samples.

## Results

### Subtype transitions and distribution across trimesters

The classification of GDM participants at T2 and T3 identified T2D-like subtypes: SIDD, SIRD, MOD, and MARD. The distribution of participants at both time points was comparable across all subtypes (Figure 1A). Most individuals were classified into the MOD and MARD T2D-like subtypes, whereas fewer were grouped into the SIDD and SIRD T2D-like subtypes. The transitions between subtypes from T2 to T3 showed that the majority of MOD and MARD remained in the same subtype, indicating overall stability in cluster assignments (Figure 1B). Notably, all individuals classified as SIRD in T3 were originally classified as MARD in T2. To further investigate this transition, we separated the samples into stable and switching samples. In addition, we showed the mean difference for each clinical variable between T2 and T3 across all subtype changes. We observed an increase in BMI from T2 to T3 across all samples except those that changed from MOD to MARD, as indicated by the reduction in BMI (Supplementary Table 1). HbA1C did not show a significant difference across subtype transitions between T2 and T3 (Supplementary Table 2). However, only the MOD subtype showed a statistically significant increase in HbA1C (*p-value* = 0.02). Interestingly, individuals classified as MARD at T2 showed a significant increase in HOMA2-IR (*p-value* = 0.04) when individuals transitioned to the SIRD subtype (Supplementary Table 3). The increase in HOMA2-IR in subtype change from MARD to SIRD was the highest among all subtypes (*p-value = 0.04)*. In contrast, the subtype change from SIRD to MARD was associated with a decrease in HOMA2-IR, but this change was not statistically significant. Similarly, the transition from MARD to SIRD showed a significant increase in HOMA2-%B (*p-value = 0.04*), whereas the SIRD transition to MARD showed a reduction in HOMA2-%B (Supplementary Table 4).

**Figure 1.**
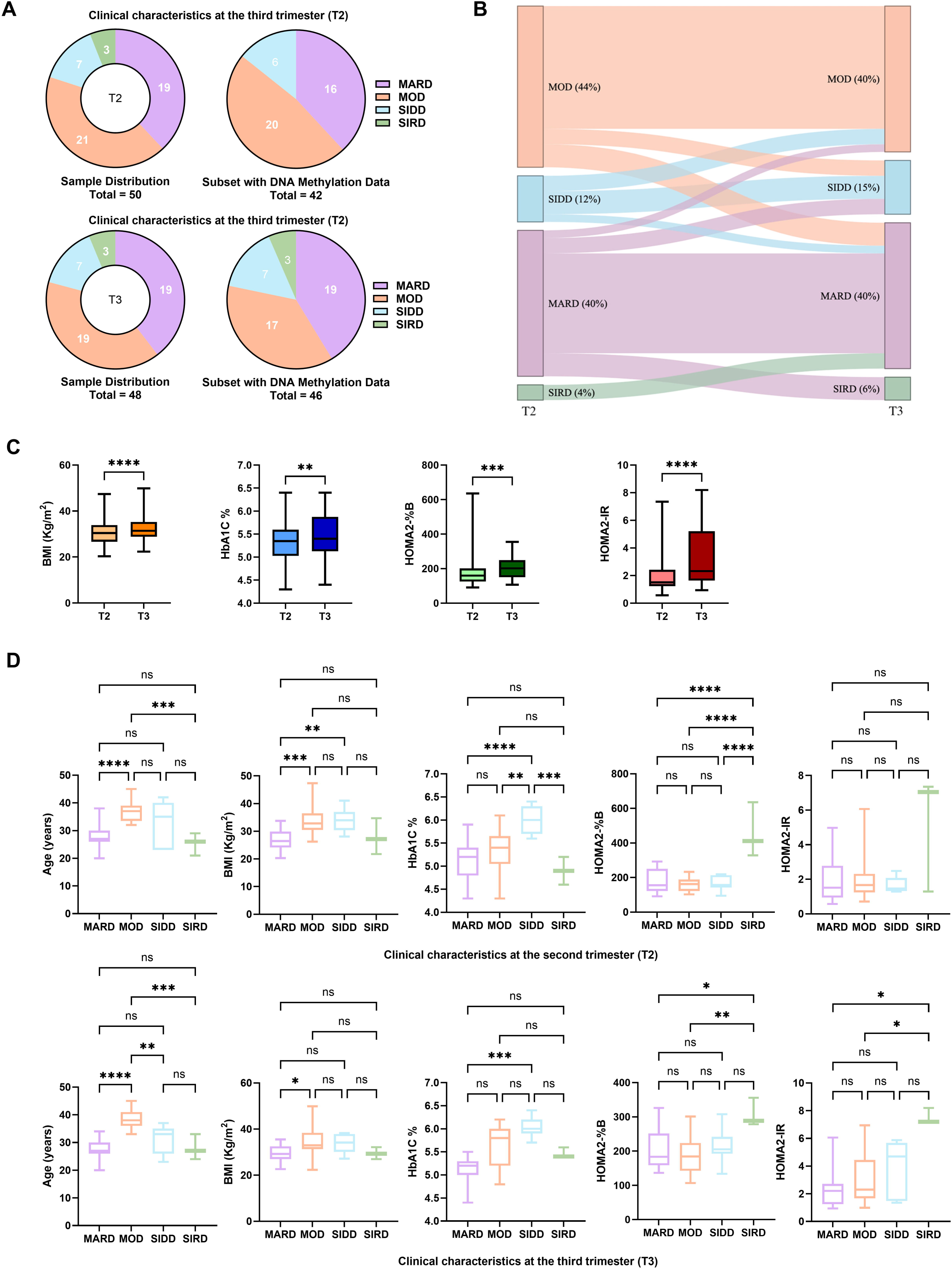
The distribution and clinical characteristics of GDM T2D-like subtypes in the QBiC cohort. A) Overall, the distribution of the GDM T2D-like subtype within the cohort and the DNA methylation data among GDM participants in T2 and T3. B) GDM T2D-like subtypes transition between the T2 and T3 trimesters. C) Distribution of clinical variables (BMI, HbA1C, HOMA2-%B, and HOMA2-IR) between T2 and T3 for GDM individuals. D) Distribution of clinical variables (Age, BMI, HbA1C, HOMA2-%B, and HOMA2-IR) between T2 and T3 for GDM T2D-like subtypes. The x-axis shows T2, T3 (second and third trimesters), or T2D-like subtypes in GDM (MARD, MOD, SIDD, SIRD), while the y-axis displays clinical variables such as Age, BMI, HbA1C, HOMA2-%B, and HOMA2-IR. Significant level represents *p-value*: * < 0.05; ** < 0.01; *** <0.001; **** < 0.0001.

### Clinical heterogeneity within GDM participants

Clinical characteristics of the GDM participants were compared across trimesters as a whole group (Figure 1C) and across T2D-like subtypes (SIDD, SIRD, MOD, and MARD) for T2 and T3 (Figure 1D). Significant differences were observed in BMI, HbA1C, HOMA2-%B, and HOMA2-IR, with T3 showing significantly higher values than T2 across all parameters (Figure 1C). Interestingly, individuals classified into the MOD subtype showed significantly higher age than the MARD and SIRD subtypes at T2, whereas the MOD subtype showed a significantly higher age than all other subtypes at T3. The BMI was significantly higher in MOD than in MARD in both trimesters. The SIDD subgroup had the highest HbA1C levels across all subgroups in T2 and remained significantly higher than MARD in T3. HOMA2-%B was significantly higher in the SIRD subtype at T2; however, at T3, SIRD remained significantly higher than MOD and MARD. Although SIRD individuals showed higher HOMA2-IR, the difference was not statistically significant at T2 due to the small sample size in this group; however, at T3, SIRD had significantly higher HOMA2-IR than MOD and MARD.

### Molecular heterogeneity within GDM samples

Our analysis identified the top combined-rank methylated sites associated with SIDD, MOD, and MARD subtypes in GDM. However, none of these sites were significant after FDR adjustment. Therefore, only the CpG sites that were consistent across the two trimesters (T2 and T3) for each subtype were considered for downstream analysis. CpG sites identified in two or more subtypes were excluded from further analysis. As a result, 1,025, 719, and 373 CpG sites were associated with SIDD, MOD, and MARD, respectively. However, no regions were identified in association with the T2D-like subtypes, because none reached statistical significance after FDR correction. A few pathways with nominal *p-values* <0.05 were identified in the KEGG pathway enrichment analysis of top-ranked CpG sites; however, none remained FDR-significant. The KEGG pathway analysis identified PPAR signaling and signaling pathways regulating pluripotency of stem cells associated with MARD. In addition, the ABC transporters signaling pathway was enriched from MOD-associated sites, while maturity-onset diabetes of the young (MODY) and sphingolipid metabolism were enriched at SIDD-specific sites (Supplementary Table 5). Gene Ontology (GO) enrichment analysis was similarly conducted and revealed one biological pathway, negative regulation of fatty acid oxidation, associated with MARD, with an FDR < 0.1 (Supplementary Table 6).

### Exploring Shared DNA Methylation Signatures Between GDM and T2D

To identify common DNA Methylation Signatures in both GDM T2D-like subtypes and T2D subtypes, a differential methylation analysis was conducted on a subset of T2D subtype individuals matched to GDM T2D-like subtype individuals. This analysis identified 11 common sites across all comparisons, representing around 0.52% of the total CpGs associated with GDM subtypes (Supplementary Table 7). Of those, only three were common within the same subgroup associated with MOD (Supplementary Table 7). Similarly, 18 CpG sites (∼0.85%) were shared with the subgroup DNA methylation sites reported by Schrader *et al.* [21], of which four were common to the MOD subtype (Supplementary Table 8).

### Epigenetic age acceleration in specific physiological systems within a particular subtype

Although no statistically significant differences in EAA were detected between subgroups, including phenoAge, GrimAge, and DunedinPACE, a consistent trend was observed in DunedinPACE (Supplementary Figures S2 and S3). DunedinPACE showed that SIDD has a higher pace of aging than MOD and MARD in T2 and T3. The system-specific age acceleration across subtypes was not statistically significant except in the kidney (Supplementary Figure S1). However, the low epigenetic age acceleration was observed at T2 only in the SIDD subgroup (Supplementary Figure S1). In addition, a consistent trend was observed in the MARD subgroup, which showed higher EAA in the brain and blood than in other systems in both trimesters (Figure 2). Similarly, SIDD showed increased age acceleration in the lung, liver, metabolic, and inflammatory systems. Age acceleration in the lung was detected in the SIRD subgroup at T3, but not at T2, due to the absence of DNA methylation data (Figure 2).

**Figure 2.**
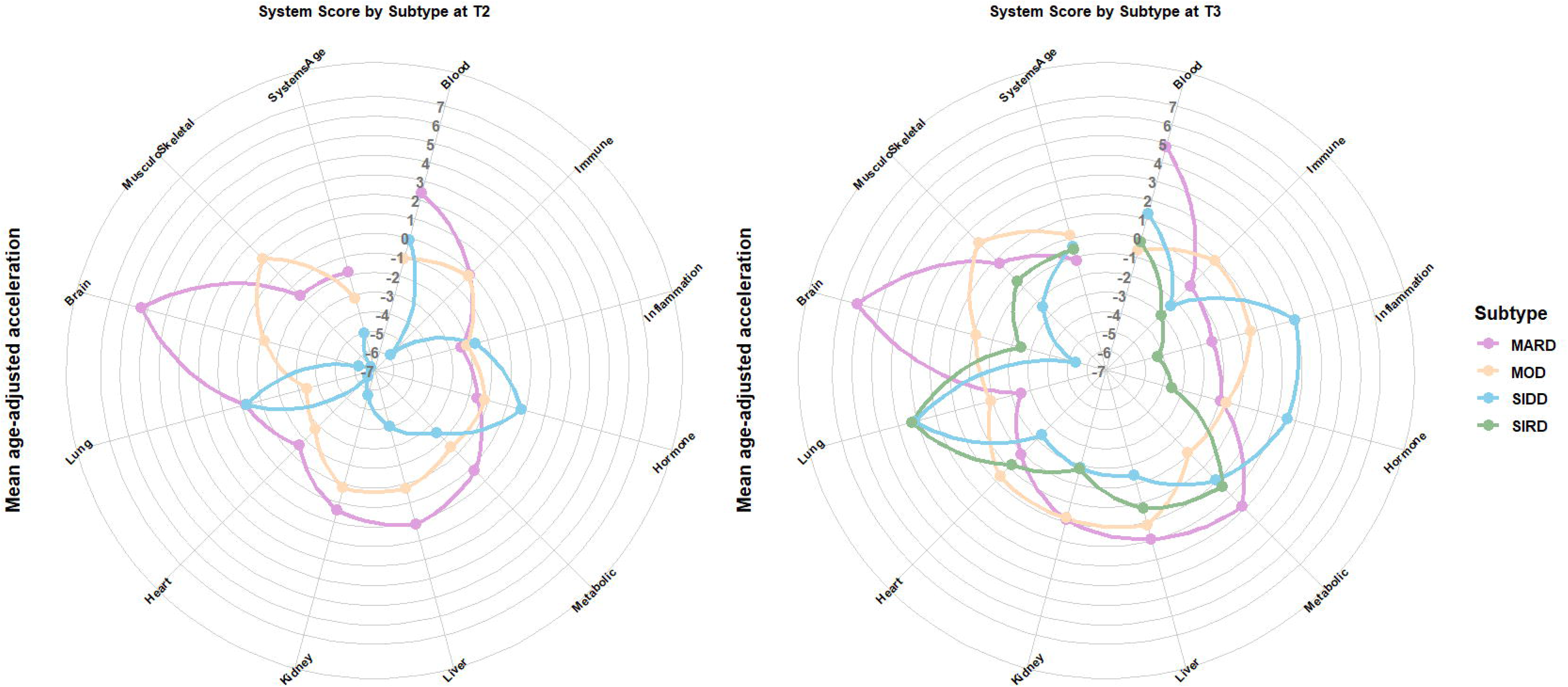
Age-adjusted acceleration (mean) across 12 physiological systems in different GDM T2D-like subtypes for T2 and T3 trimesters. The spider plots show circles on a standardized scale, with each ring representing a 1-unit increase in age acceleration. Values near zero indicate alignment between biological and chronological age; higher (positive) values reflect accelerated biological aging relative to chronological age, and lower (negative) values indicate decelerated biological aging.

## Discussion

For the first time, we showed the applicability of the Ahlqvist *et al.* [7] clustering scheme on GDM. We identified GDM subgroups corresponding to the four T2D-like subtypes (SIDD, SIRD, MOD, and MARD). GDM was previously classified into insulin-deficient GDM, insulin-resistant GDM, mixed GDM (defects in both insulin secretion and sensitivity), and unclassified GDM (no defects in insulin secretion or sensitivity) [22]. However, their classification relied on a different approach, using indices from the oral glucose tolerance test to estimate insulin secretion and sensitivity, yielding distinct outcomes [22]. The longitudinal data used in our study revealed short-term subtype transitions during pregnancy while maintaining a comparable subtype distribution across trimesters. The distribution of GDM subtypes was consistent with previous reports on T2D in the Qatari population, with most samples clustering in MOD and MARD [18, 23]. Distinct clinical characteristics were also observed across GDM subtypes in our study. These characteristics were comparable to those described in earlier studies of T2D subgroups [18, 23]. There is evidence of increasing insulin resistance in GDM across all trimesters [Thilak, 2023 #70], which may explain the T2D-like subtype changes from SIRD to MARD (T2) and from MARD to SIRD (T3) due to elevated HOMA2-IR levels in those individuals.

We identified subtype-specific epigenetic signatures that highlight molecular heterogeneity and further support the biological distinction between GDM T2D-like subtypes. Distinct epigenetic alterations were observed in GDM, as reflected by the low overlap in CpG sites identified when exploring shared DNA methylation signatures between GDM and T2D. Nevertheless, the potentially dysregulated pathways identified suggested overlapping molecular mechanisms underlying both conditions. These pathways included the PPAR signaling pathway, which plays a role in lipid metabolism, glucose homeostasis, and insulin resistance [24]. Furthermore, PPAR is implicated in maternal adaptation during pregnancy, as well as in GDM [25]. Targeting PPAR nuclear receptors with agonists has been reported in T2D; however, safety concerns and limited knowledge prevent its use in GDM treatment. [25]. Sphingolipid metabolism was also identified among the enriched pathways in SIDD. Furthermore, research in the Qatari population demonstrated that sphingolipids were downregulated in T2D SIDD [23]. Early postpartum metabolomics in women with GDM showed significant downregulation of sphingolipid metabolism, which is linked to the transition from GDM to T2D [26]. Additional studies are needed to investigate the role of sphingolipid metabolism during pregnancy. Furthermore, our analysis found a significant association with the biological process of negatively regulating fatty acid oxidation. Our finding was consistent with earlier reports indicating impaired free fatty acid β-oxidation in skeletal muscle cells from individuals with T2D, and that PPARγ treatment reversed these impairments [27]. This finding highlights the potential importance of PPAR signaling pathways in the pathophysiology of both GDM and T2D.

Previous studies suggest that T2D subtypes exhibit distinct profiles of complications. For instance, the SIDD subtype is associated with a higher risk of retinopathy and neuropathy, while the SIRD subtype is more frequently linked to nephropathy and fatty liver disease [7]. Due to the similarity highlighted previously between the two conditions, we investigated age acceleration at the physiological systems level. However, no significant EAA was observed across the subgroups except in the kidney, where lower epigenetic age acceleration was observed in the SIDD subgroup at T2. This observation exhibited inconsistency across the two trimesters and may be attributable to the limited sample size within that subgroup. On the other hand, a trend toward increased EAA was observed in the MARD subgroup, specifically in the brain and blood. This pattern is consistent across all trimesters, but it was not statistically significant compared to other subtypes. Overall, no significant epigenetic age acceleration was observed in any physiological system, suggesting that the T2D-like subtype-associated long-term complications previously discussed are unlikely to be detected within a short period.

We recognize several limitations in this study, including a small sample size that limited the statistical power of our analysis. Additionally, SIRD was excluded from further differential DNA methylation analysis because no DNA methylation data were available for the samples identified in the SIRD subgroup. Another limitation is the lack of postpartum follow-up data for mothers and offspring, including future maternal T2D status and fetal outcomes such as birthweight. Consequently, future development of T2D was not determined, and the predictive value of the identified T2D-like subtype-specific methylation signatures for future disease progression was not assessed.

Overall, our study applied for the first time a novel clustering method, first designed for T2D, to pregnant women with GDM. We found that most GDM participants classified into the mild T2D-like subtypes, MOD (42%) and MARD (38%). In contrast, only a small percentage represent severe cases, in T2D-like subtypes SIDD (14%) and SIRD (6%), indicating variable severity within GDM. Furthermore, we demonstrated subgroup stability across different trimesters of pregnancy T2 and T3. The limited overlap in CpGs between GDM and T2D might suggest that GDM has a distinct epigenetic landscape, despite sharing clinical and metabolic features with T2D. However, further longitudinal studies with postpartum follow-up in larger, more diverse cohorts are warranted to validate these findings and determine whether subtype classification has clinical utility for personalized monitoring, prognosis, and treatment strategies.

## Funding

This work was supported by an NPRP13 grant (NPRP13S-0113-200050) from the Qatar National Research Fund (QNRF).

## Contributions

L.S.: Writing – original draft, Writing – review & editing, and conducting experiments and data analysis. N.M.A.: Writing – original draft, Writing – review & editing, and data analysis. E.F.: Providing samples, writing, review & editing. O.M.E.A.: Supervision, Writing – review & editing. N.E.H.: Supervision, Conceptualization, Funding, Writing – review & editing. All authors have thoroughly reviewed and consented to the publication of the manuscript.

## Conflict of interest

The authors have no conflicts of interest.

## Acknowledgments

The authors would like to thank the QBiC participants, the research team, and all Qatar Biobank personnel for their contribution to this study.

## Data Availability

The datasets generated and analyzed in this study are available from the corresponding author upon reasonable request, subject to institutional and ethical approvals.

## Notes

### Competing Interest Statement

The authors have declared no competing interest.

### Author Declarations

This study was approved by the institutional review boards of QPHI (Ex-2022-QF-QBB-RES-ACC-00100-0203) and Hamad Bin Khalifa University (QBRI-IRB 2021-09-107).

