## Supplementary Table, Supplementary Figure for "Evaluation of Type 2 Diabetes Like Subtypes in Gestational Diabetes"

**Supplementary Materials**

**Table S1:** BMI change between T3 and T2 across all subtype changes.

| **GDM T2D-like Subtype change** | **N** | **Mean BMI at T2** | **Mean BMI at T3** | **BMI change** | **Test** | ***p-value*** |
| --- | --- | --- | --- | --- | --- | --- |
| MARD → MARD | 13 | 27.56 | 29.33 | 1.77 | Paired t-test | 5.80e-05 |
| MOD → MOD | 16 | 34.04 | 35.48 | 1.44 | Paired t-test | 3.91e-05 |
| MOD → SIDD | 2 | 33.91 | 36.19 | 2.28 | NA | NA |
| SIRD → MARD | 2 | 24.46 | 27.49 | 3.02 | NA | NA |
| MARD → SIDD | 2 | 24.81 | 28.71 | 3.89 | NA | NA |
| MOD → MARD | 3 | 31.21 | 31.02 | -0.19 | Paired t-test | 8.40e-01 |
| MARD → SIRD | 3 | 27.17 | 29.49 | 2.32 | Paired t-test | 4.38e-02 |
| SIDD → SIDD | 3 | 33.91 | 34.43 | 0.52 | Paired t-test | 4.78e-01 |
| SIDD → MOD | 2 | 32.60 | 32.80 | 0.20 | NA | NA |
| SIDD → MARD | 1 | 28.11 | 28.89 | 0.78 | NA | NA |
| MARD → MOD | 1 | 20.53 | 22.30 | 1.77 | NA | NA |

**Table S2:** HbA1C change between T3 and T2 across all subtype changes.

| **GDM T2D-like Subtype change** | **N** | **Mean HbA1C at T2** | **Mean HbA1C at T3** | **HbA1C change** | **Test** | ***p-value*** |
| --- | --- | --- | --- | --- | --- | --- |
| MARD → MARD | 13 | 5.08 | 5.17 | 0.092 | Paired t-test | 0.21 |
| MOD → MOD | 16 | 5.36 | 5.53 | 0.18 | Paired t-test | 0.02 |
| MOD → SIDD | 2 | 5.55 | 6.20 | 0.65 | NA | NA |
| SIRD → MARD | 2 | 4.75 | 5.10 | 0.35 | NA | NA |
| MARD → SIDD | 2 | 5.35 | 5.80 | 0.45 | NA | NA |
| MOD → MARD | 3 | 5.23 | 5.07 | -0.17 | Wilcoxon signed-rank | 0.17* |
| MARD → SIRD | 3 | 5.53 | 5.47 | -0.07 | Paired t-test | 0.80 |
| SIDD → SIDD | 3 | 6.13 | 6.13 | 0 | Paired t-test | 1.00 |
| SIDD → MOD | 2 | 6.10 | 5.90 | -0.20 | NA | NA |
| SIDD → MARD | 1 | 5.60 | 5.50 | -0.10 | NA | NA |
| MARD → MOD | 1 | 4.70 | 5.00 | 0.30 | NA | NA |

* The Wilcoxon signed-rank test was used due to the non-normal distribution; p-values were calculated using this non-parametric test, while mean values are shown descriptively.

**Table S3:** HOMA2-IR change between T3 and T2 across all subtype changes.

| **GDM T2D-like Subtype change** | **N** | **Mean HOMA2-IR at T2** | **Mean HOMA2-IR at T3** | **HOMA2-IR change** | **Test** | ***p-value*** |
| --- | --- | --- | --- | --- | --- | --- |
| MARD → MARD | 13 | 1.93 | 2.57 | 0.64 | Paired t-test | 0.16 |
| MOD → MOD | 16 | 1.91 | 3.28 | 1.37 | Wilcoxon signed-rank | 0.02* |
| MOD → SIDD | 2 | 3.95 | 5.19 | 1.24 | NA | NA |
| SIRD → MARD | 2 | 4.31 | 2.72 | -1.60 | NA | NA |
| MARD → SIDD | 2 | 0.78 | 3.48 | 2.70 | NA | NA |
| MOD → MARD | 3 | 1.46 | 2.87 | 1.41 | Paired t-test | 0.05 |
| MARD → SIRD | 3 | 3.50 | 7.51 | 4.01 | Paired t-test | 0.04 |
| SIDD → SIDD | 3 | 1.56 | 3.55 | 1.98 | Paired t-test | 0.26 |
| SIDD → MOD | 2 | 1.90 | 1.74 | -0.16 | NA | NA |
| SIDD → MARD | 1 | 1.41 | 0.94 | -0.48 | NA | NA |
| MARD → MOD | 1 | 0.93 | 2.62 | 1.70 | NA | NA |

* The Wilcoxon signed-rank test was used due to the non-normal distribution; p-values were calculated using this non-parametric test, while mean values are shown descriptively.

**Table S4:** HOMA2%B change between T3 and T2 across all subtype changes.

| **GDM T2D-like Subtype change** | **N** | **Mean HOMA2%B at T2** | **Mean HOMA2%B at T3** | **HOMA2%B change** | **Test** | ***p-value*** |
| --- | --- | --- | --- | --- | --- | --- |
| MARD → MARD | 13 | 184.06 | 206.92 | 22.85 | Wilcoxon signed-rank | 0.26* |
| MOD → MOD | 16 | 153.06 | 182.86 | 29.81 | Paired t-test | 0.01 |
| MOD → SIDD | 2 | 198.65 | 223.55 | 24.90 | NA | NA |
| SIRD → MARD | 2 | 523.50 | 276.95 | -246.55 | NA | NA |
| MARD → SIDD | 2 | 138.50 | 209.35 | 70.85 | NA | NA |
| MOD → MARD | 3 | 167.93 | 194.53 | 26.60 | Paired t-test | 0.17 |
| MARD → SIRD | 3 | 183.23 | 307.63 | 124.40 | Paired t-test | 0.04 |
| SIDD → SIDD | 3 | 149.13 | 213.33 | 64.20 | Paired t-test | 0.33 |
| SIDD → MOD | 2 | 153.90 | 177.20 | 23.30 | NA | NA |
| SIDD → MARD | 1 | 167.60 | 148.40 | -19.20 | NA | NA |
| MARD → MOD | 1 | 140.00 | 300.50 | 160.50 | NA | NA |

* The Wilcoxon signed-rank test was used due to the non-normal distribution; p-values were calculated using this non-parametric test, while mean values are shown descriptively.

**Table S5:** KEGG pathways enriched from CpG sites identified in the GDM subtype.

| **KEGG pathway** | **Description** | **N** | **DE** | **P.DE** | **FDR** | **GDM T2D-like Subtypes** |
| --- | --- | --- | --- | --- | --- | --- |
| hsa04550 | Signaling pathways regulating pluripotency of stem cells | 143 | 3 | 0.006116 | 1 | MARD |
| hsa03320 | PPAR signaling pathway | 74 | 2 | 0.01181 | 1 | MARD |
| hsa02010 | ABC transporters | 45 | 2 | 0.009706 | 1 | MOD |
| hsa00511 | Other glycan degradation | 18 | 2 | 0.003195 | 1 | SIDD |
| hsa04950 | Maturity-onset diabetes of the young | 26 | 2 | 0.006947 | 1 | SIDD |
| hsa00600 | Sphingolipid metabolism | 55 | 2 | 0.02574 | 1 | SIDD |
| hsa00440 | Phosphonate and phosphinate metabolism | 6 | 1 | 0.02788 | 1 | SIDD |

**Table S6:** Gene ontology terms enriched among CpG sites identified in the MARD subtype.

|  | **Ontology** | **Term** | **N** | **DE** | **P.DE** | **FDR** |
| --- | --- | --- | --- | --- | --- | --- |
| GO:0046322 | BP | negative regulation of fatty acid oxidation | 14 | 3 | 4.07x 10^-06^ | 0.09173 |

**Table S7:** Overlapping CpG sites identified in GDM-like T2D subtypes with T2D subtypes.

| **CpG** | **Chr** | **Gene Symbol** | **Gene Region** | **CpG Island Group** | **T2D Subtypes** | **GDM T2D-like Subtypes** |
| --- | --- | --- | --- | --- | --- | --- |
| cg21122483 | 16 |  |  | OpenSea | MOD | MOD |
| cg25854064 | 4 |  |  | OpenSea | MOD | MOD |
| cg08587775 | 19 |  |  | Island | MOD | MARD |
| cg06211550 | 16 |  |  | N_Shelf | SIDD | MOD |
| cg15440376 | 4 |  |  | OpenSea | SIDD | MOD |
| cg20069666 | 14 |  |  | OpenSea | SIDD | MOD |
| cg00951857 | 6 |  |  | N_Shore | SIRD | MARD |
| cg15351009 | 5 | LRRC14B | TSS200 | N_Shore | SIRD | MARD |
| cg06313716 | 4 |  |  | Island | MOD | MOD |
| cg19335003 | 5 | PCDHA4 | TSS1500 | N_Shore | MOD | SIDD |
| cg02094681 | 5 |  |  | OpenSea | SIRD | MOD |

**Table S8:** Look-up of CpGs previously identified as subgroup-unique DNA methylation sites in T2D by Schrader *et al*.

| **CpG** | **Chr** | **Gene Symbol** | **Gene Region** | **CpG Island Group** | **T2D Subtypes** | **GDM T2D-like Subtypes** |
| --- | --- | --- | --- | --- | --- | --- |
| cg22891868 | 2 | *MOGAT1* | TSS1500 | N_Shore | SIDD | MOD |
| cg18645241 | 21 |  | intergenic | open_sea | MOD | MARD |
| cg08986196 | 2 | *FAM228B;PFN4* | 5'UTR;TSS1500;Body | S_Shore | MOD | MARD |
| cg05616442 | 14 | *AKT1* | 5'UTR;TSS1500 | N_Shore | MOD | MARD |
| cg02905815 | 13 |  | intergenic | open_sea | MOD | MARD |
| cg13379325 | 20 | *KCNQ2* | Body | Island | SIDD | MOD |
| cg14484690 | 6 | *LINC00271* | Body | open_sea | MOD | MOD |
| cg10832308 | 13 | *ENOX1* | Body | open_sea | MOD | MOD |
| cg07822788 | 6 | *NOTCH4* | TSS1500 | open_sea | MOD | MOD |
| cg20131013 | 17 | *RICH2* | TSS1500 | N_Shore | MOD | MOD |
| cg17233022 | 1 | *SESN2* | TSS1500 | N_Shore | MOD | SIDD |
| cg18681014 | 14 |  | intergenic | N_Shore | MOD | SIDD |
| cg20224311 | 12 | *CCT2* | TSS1500 | N_Shore | MOD | SIDD |
| cg22891191 | 10 | *PTPRE* | 5'UTR | Island | MOD | SIDD |
| cg21188652 | 2 | *RQCD1* | Body | S_Shore | MOD | SIDD |
| cg09558502 | 1 | *OVGP1* | TSS200 | open_sea | MOD | SIDD |
| cg03316864 | 19 | *B3GNT3* | 5'UTR | Island | MOD | SIDD |
| cg02828862 | 9 | *PTPRD* | 5'UTR | open_sea | MOD | SIDD |

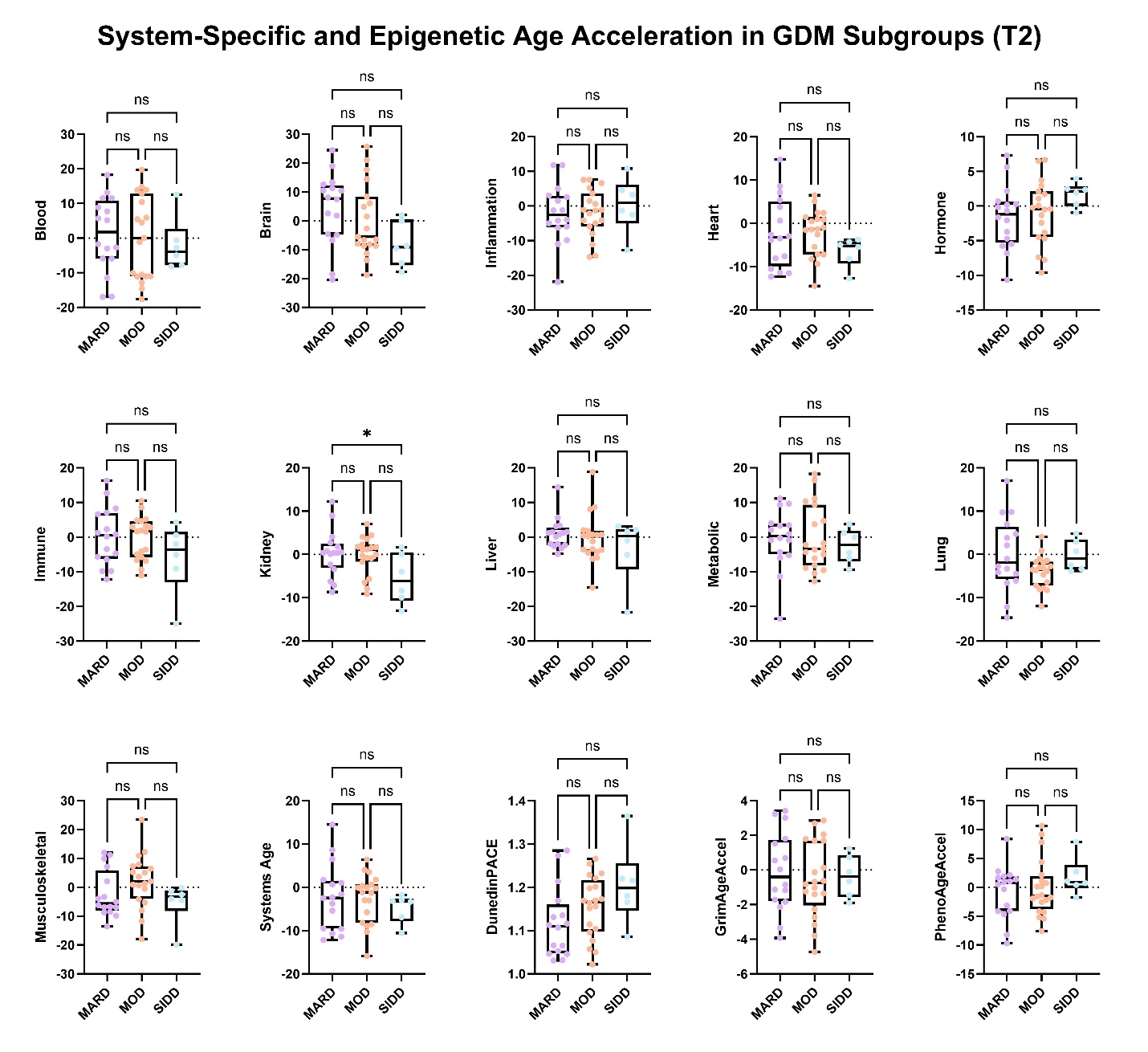

**Figure S1:** Boxplots showing the comparison of age acceleration across multiple physiological systems (heart, lung, kidney, liver, brain, immune, inflammatory, blood, musculoskeletal, hormone, and metabolic) and epigenetic age acceleration measured using DunedinPACE, GrimAge, and PhenoAge clocks across GDM subtypes, including MARD, MOD, and SIDD, at T2. Each boxplot displays the median and interquartile range (IQR), while the whiskers indicate the minimum and maximum values. Significant level represents *p-value*: * < 0.05; ** < 0.01; *** <0.001; **** < 0.0001.

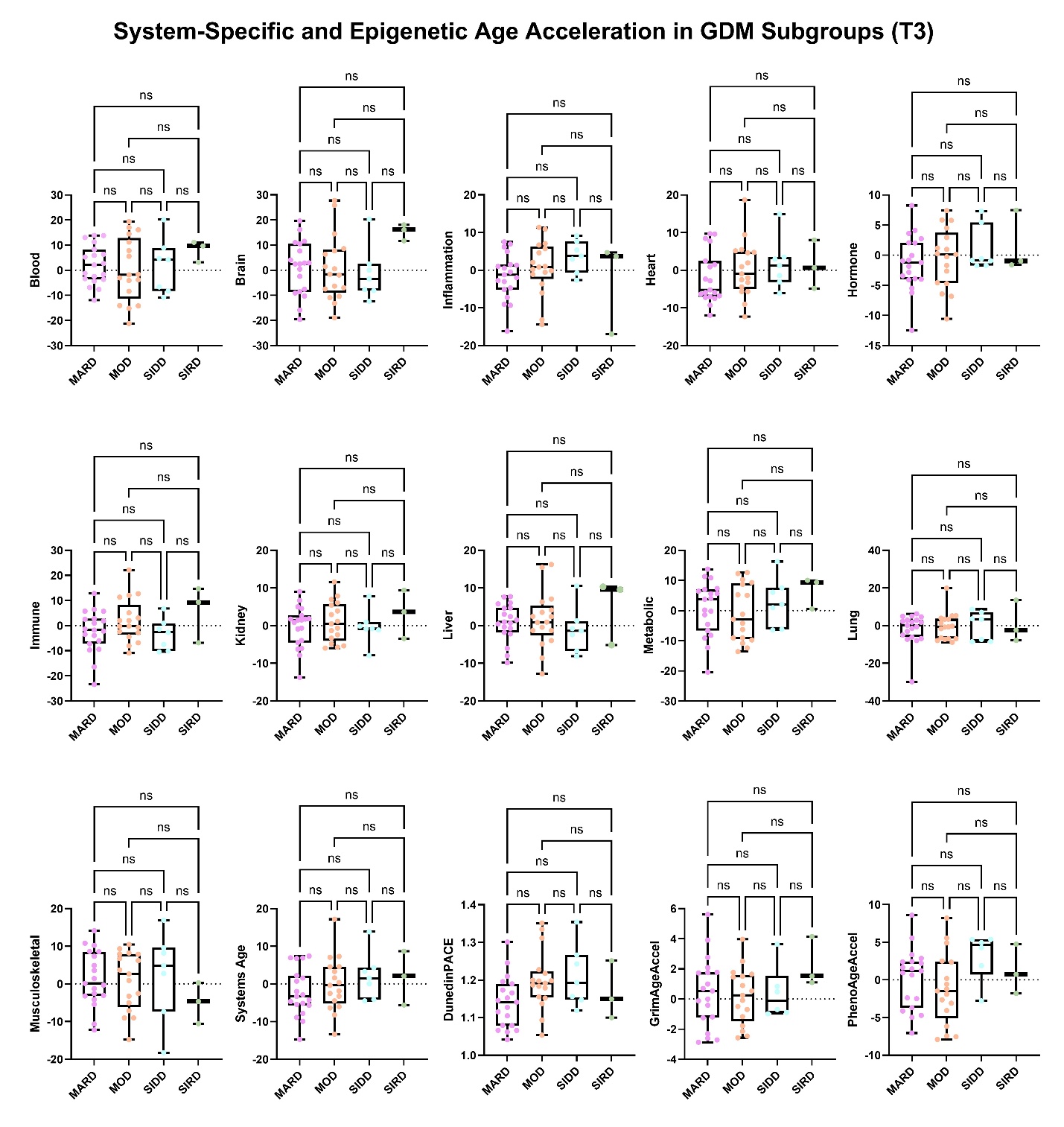

**Figure S2:** Boxplots showing the comparison of age acceleration across multiple physiological systems (heart, lung, kidney, liver, brain, immune, inflammatory, blood, musculoskeletal, hormone, and metabolic) and epigenetic age acceleration measured using DunedinPACE, GrimAge, and PhenoAge clocks across GDM subtypes, including MARD, MOD, SIDD, and SIRD at T3. Each boxplot displays the median and interquartile range (IQR), while the whiskers indicate the minimum and maximum values. Significant level represents *p-value*: * < 0.05; ** < 0.01; *** <0.001; **** < 0.0001.
